# Assessing the clinical application of deep-learning-derived CT volumetric measures in neurodegenerative disease diagnostics

**DOI:** 10.1101/2022.12.19.22282455

**Authors:** Meera Srikrishna, Nicholas J. Ashton, Alexis Moscoso, Joana B. Pereira, Rolf A. Heckemann, Danielle van Westen, Giovanni Volpe, Joel Simrén, Anna Zettergren, Silke Kern, Lars-Olof Wahlund, Bibek Gyanwali, Saima Hilal, Joyce Chong Ruifen, Henrik Zetterberg, Kaj Blennow, Eric Westman, Christopher Chen, Ingmar Skoog, Michael Schöll

**Affiliations:** Wallenberg Centre for Molecular and Translational Medicine, University of Gothenburg, Gothenburg, Sweden; Department of Psychiatry and Neurochemistry, Institute of Physiology and Neuroscience, University of Gothenburg, Gothenburg, Sweden; King’s College London, Institute of Psychiatry, Psychology and Neuroscience, Maurice Wohl Institute Clinical Neuroscience Institute, London, UK; NIHR Biomedical Research Centre for Mental Health and Biomedical Research Unit for Dementia at South London and Maudsley NHS Foundation, London, UK; Division of Clinical Geriatrics, Department of Neurobiology, Care Sciences and Society, Karolinska Institutet, Stockholm, Sweden; Memory Research Unit, Department of Clinical Sciences, Malmö Lund University, Malmö, Sweden; Department of Medical Radiation Sciences, Institute of Clinical Sciences, Sahlgrenska Academy, Göteborg, Sweden; Department of Clinical Sciences, Diagnostic Radiology, Lund University Sweden; Department of Imaging and Function, Skånes University Hospital, Lund, Sweden; Department of Physics, University of Gothenburg, SE-41296 Gothenburg, Sweden; Clinical Neurochemistry Laboratory, Sahlgrenska University Hospital, Mölndal, Sweden; Neuropsychiatric Epidemiology, Institute of Neuroscience and Physiology, Sahlgrenska Academy, Centre for Ageing and Health (AgeCap), University of Gothenburg, Gothenburg, Sweden; Department of Psychiatry and Neurochemistry, Institute of Neuroscience and Physiology, Sahlgrenska Academy, University of Gothenburg, Mölndal, Sweden; Psychiatry Cognition and Old Age Psychiatry Clinic, Sahlgrenska University Hospital, Region Västra Götaland, Sweden; Memory Aging and Cognition Centre, National University Health System, Singapore; Department of Pharmacology, Yong Loo Lin School of Medicine, National University of Singapore, Singapore; Saw Swee Hock School of Public Health, National University of Singapore and National University Health System, Singapore; Department of Neurodegenerative Disease, UCL Institute of Neurology, London, UK; UK Dementia Research Institute at UCL, London, UK; Hong Kong Center for Neurodegenerative Diseases, Hong Kong, China; Wisconsin Alzheimer’s Disease Research Center, University of Wisconsin School of Medicine and Public Health, University of Wisconsin-Madison, Madison, WI, USA; Dementia Research Centre, Institute of Neurology, University College London, London, UK; Department of Clinical Physiology, Sahlgrenska University Hospital, Gothenburg, Sweden

**Keywords:** CT: MRI, Alzheimer’s disease, vascular dementia, brain atrophy, deep learning

## Abstract

**Background:** Existing bio fluid and imaging biomarkers used in research and clinical diagnostics of neurodegenerative diseases are often expensive or invasive and are mainly available in specialised care centres. CT is an affordable and widely available imaging modality predominantly used to evaluate structural abnormalities, but not for the volumetric quantification of neurodegeneration. Previously, we developed a deep learning model trained on MRI segmentations from individuals with paired CT and MR scans, which achieved high accuracy and robust tissue classification based on brain CT images.

**Purpose:** To explore the diagnostic utility of deep-learning-derived CT-based atrophy measures and study their association with relevant cognitive, biochemical and other imaging markers of neurodegenerative diseases.

**Materials and methods:** In this retrospective study, we analysed 917 CT and 744 MR scans from cognitively healthy participants of the Gothenburg H70 Birth Cohort (70.4 ± 2.6 years) and 204 CT and 241 MR scans from participants of the Memory Clinic Cohort, Singapore (73 Alzheimer’s disease, 20 vascular dementia, 22 cognitively normal; 74.0 ± 8.2 years). We tested associations between six CT-derived volumetric measures with clinical diagnosis, fluid and imaging biomarkers and cognition.

**Results:** In the Memory Clinic Cohort, deep-learning-derived CT-based atrophy measures differentiated cognitively healthy individuals from Alzheimer’s disease (AUC 0.88; 95% CI: 0.79-0.96) and vascular dementia (AUC 0.91; 95% CI: 0.81-1.00) patients with high accuracy levels comparable to MR-derived measures. Additionally, CT-based measures distinguished early, prodromal Alzheimer’s disease (AUC= 0.73, 95% CI: 0.62, 0.85) and prodromal vascular dementia patients from healthy individuals (CT-GM: AUC= 0.7, 95% CI: 0.51, 0.81). CT-derived volumes were significantly associated with measures of cognition and biochemical markers of neurodegeneration, notably plasma-derived neurofilament light (ρ=-0.43, p<0.001, in the Memory Clinic Cohort).

**Conclusion:** Our findings provide strong evidence for the potential of deep-learning-derived CT-based atrophy measures in aiding neurodegenerative disease diagnostics in primary care settings.

## 1. Introduction

Neurodegenerative diseases are characterised by a progressive loss of neuronal integrity and function as well as molecular abnormalities leading to cognitive decline and dementia^1,2^. The most established biomarkers for neurodegenerative diseases include cerebrospinal fluid (CSF) measures of neuronal injury^3^; brain atrophy measured on structural MRI^4,5^, and PET imaging of brain glucose metabolism, as well as disease-specific measures of amyloid-β (Aβ), and tau^6,7^. However, these methods are either expensive, invasive, or available only in specialised care centres. Brain imaging modalities such as CT and recently established blood plasma-derived biomarkers^8,9^ can address the issue of cost and availability to some extent. With increased diagnostic utility, CT and plasma biomarkers may become scalable and economical alternatives for neurodegenerative disease diagnostics.

CT is a fast, low-cost and widely available imaging modality and an alternative for patients who cannot undergo an MRI examination^10^. CT-based visual ratings are comparable to those obtained from MRI with regard to certain pathomorphological characteristics, and they are significantly correlated with cognitive test results^10,11^. However, MRI is extensively used for high-resolution atrophy assessment and brain volumetry^12–14^ due to its stronger soft-tissue contrast. Atrophy assessment in CT is conducted through semi-quantitative visual ratings, a subjective, time-consuming approach that requires a trained expert. Recent studies have explored state-of-the-art techniques such as deep learning in automatic brain CT segmentation^15,16^. We have previously conducted a study where we used MR-derived tissue class segmentations to train deep learning models to perform tissue classification on brain CT images^17^. In that study, U-Net^18^ deep learning models were trained to segment grey matter (GM), white matter (WM), CSF, and intracranial volume (ICV) in brain CT images. While the segmentation results agreed strongly with those of the reference methods, the question of clinical utility was relegated to future work.

In the present study, we assessed the clinical utility of CT-based tissue classification by 1) deriving volumetric measures from CT-based tissue class maps, 2) evaluating the diagnostic value of these measures in distinguishing patients with neurodegenerative disorders from healthy study participants, and 3) exploring the relationship between these measures and cognition, as well as relevant blood and CSF biomarkers. The results will help to assess the potential of automatic CT image analysis, along with other easily accessible and scalable diagnostic tools such as blood biomarkers, for clinical diagnostic applications. Figure 1 describes the potential application of deep-learning-derived CT-based measures as a first-line investigation to support and improve diagnostics in neurodegenerative diseases, specifically dementia disorder.

**Figure 1.**
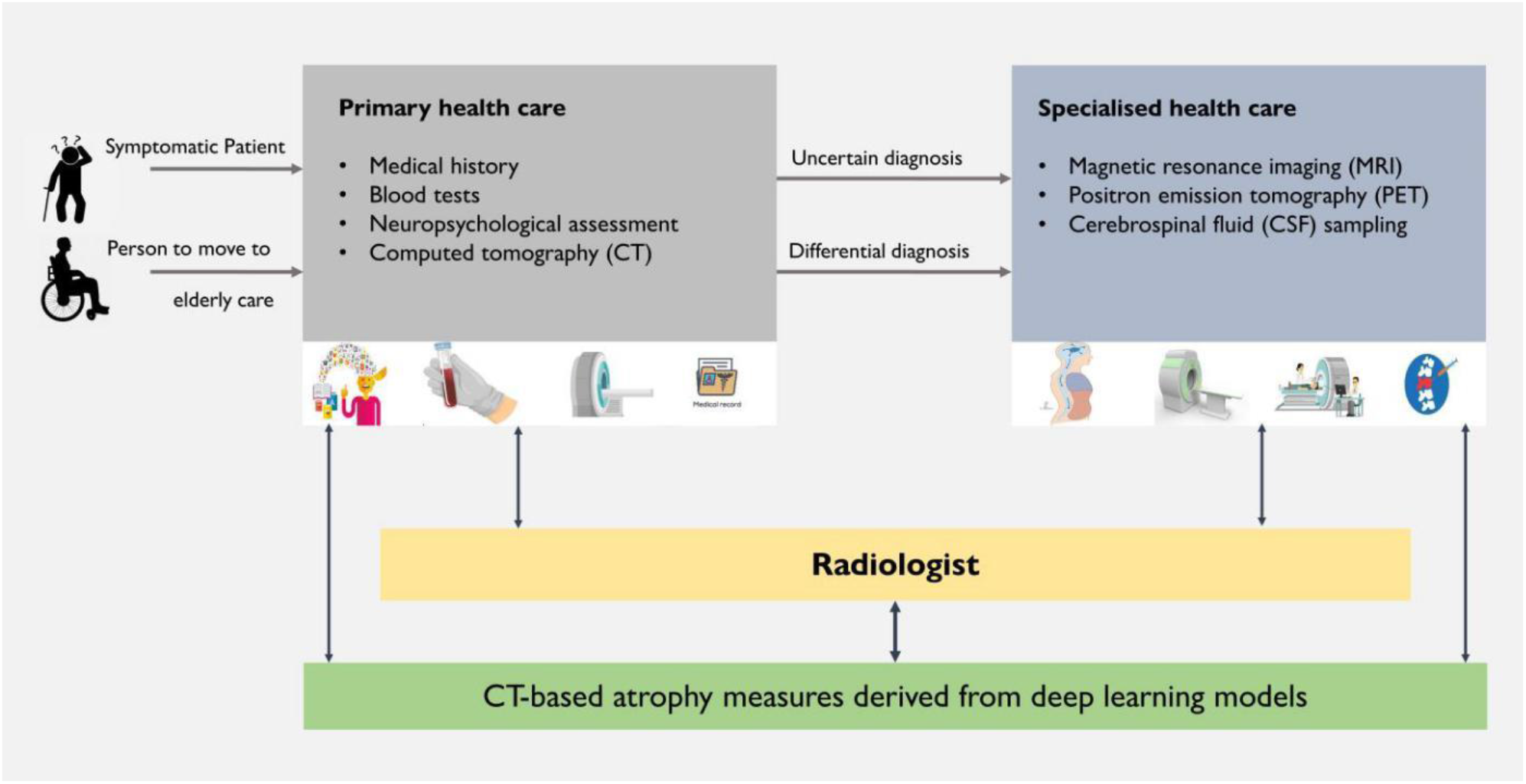
Potential clinical application of deep-learning-derived CT-based measures in neurodegenerative disease diagnostics. When a symptomatic individual enrolls in primary health care, along with blood tests, CT examinations are routinely conducted in this stage mainly to visually assess brain integrity and identify strokes, tumours, and other structural abnormalities. In case of further or uncertain diagnosis, the individual is referred to a specialised care centre where an MRI, PET or cerebrospinal fluid sampling is conducted. In general clinical settings, CT is a first-line assessment tool and the objective of this study was to automatically extract brain atrophy-related volumetric measures in order to support decisions on which patients should undergo more expensive/invasive specialised tests.

## 2. Materials and Methods

### Datasets

#### Gothenburg H70 Birth Cohort

The Gothenburg H70 Birth Cohort studies are a series of epidemiological investigations carried out periodically since 1971 on large, representative samples of those residents of Gothenburg, Sweden who turn 70 during a particular period^19^. We used data from the 2014–2016 iteration, which included *n*=1203 participants in total. CT images were available for *n*=917 participants (Figure 2). As expected, a large majority of these (99%; *n*=904) were cognitively normal.

**Figure 2.**
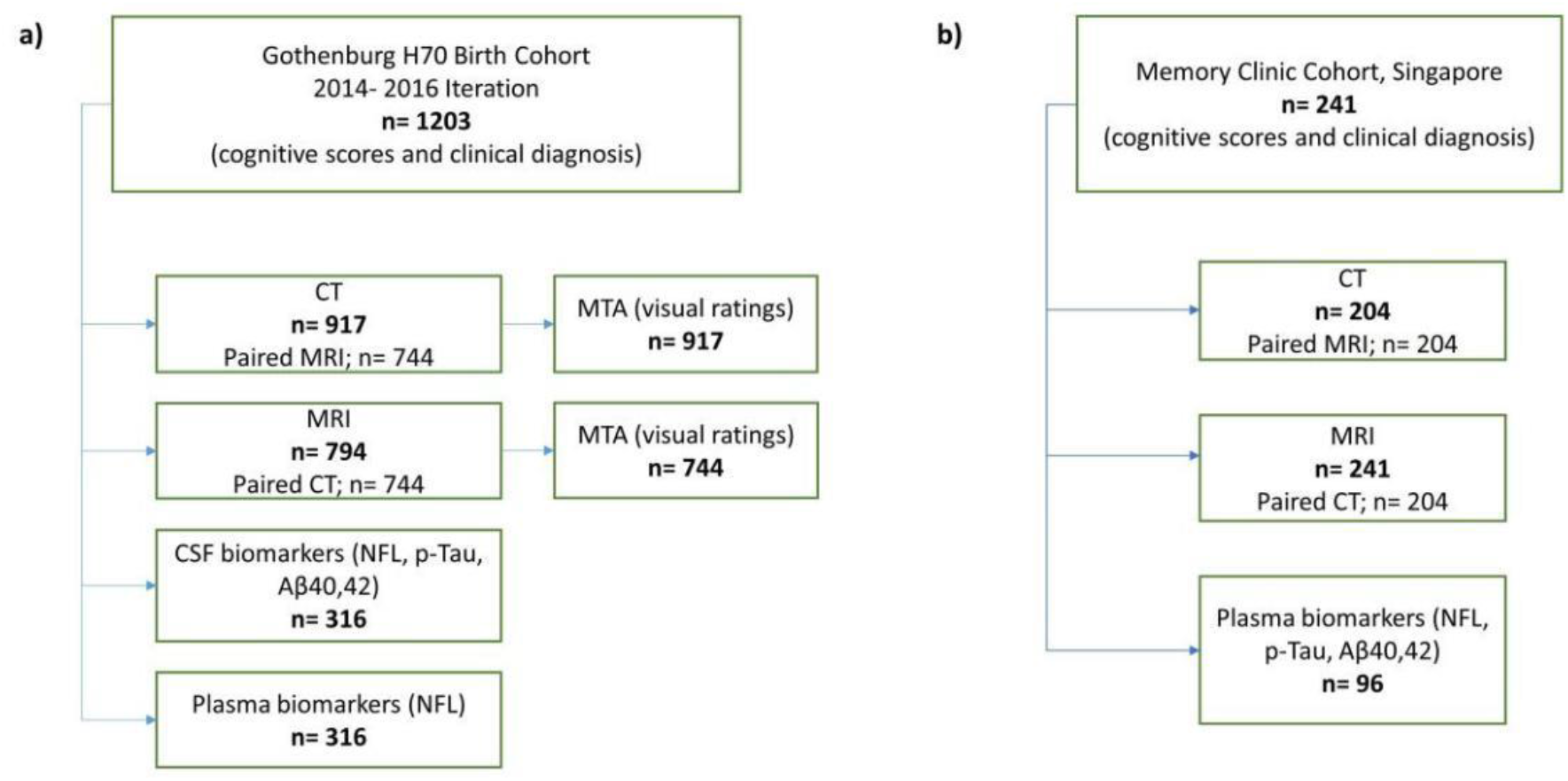
Flowchart showing the details of a) Gothenburg H70 Birth Cohort and b) Memory Clinic Cohort, Singapore. Abbreviations: Aβ, amyloid beta; CSF, cerebrospinal fluid; CT, computed tomography; MRI, magnetic resonance imaging; MTA, medial temporal atrophy; NFL, neurofilament light; p-tau, phosphorylated tau

Of these 917 participants, 79% (*n*=744) underwent MR scans within a day of CT scanning. All imaging examinations were conducted at Aleris Röntgen Annedal imaging centre in Gothenburg (Aleris Group AB, Stockholm, Sweden). A 64-slice Ingenuity CT system (Philips Medical Systems, Best, Netherlands) was used for CT acquisition, and an Achieva system (3 Tesla; Philips Medical Systems) was used for MR imaging. Informed consent had been obtained from the participants, and safety procedures were conducted through interviews. An experienced radiologist performed visual ratings of medial temporal atrophy (MTA) on all available CT and MR scans.

Demographic data, past medical history, and neuropsychological assessments were also obtained from all included individuals. CSF biomarkers of neurodegeneration and AD pathophysiology (neurofilament light (NfL), p-Tau (phosphorylated-tau), Aβ40, and Aβ42) were available for 34% (*n*=316) of participants with CT. Details of CSF sampling and analyses are provided in Rydberg Sterner et al.^19–21^. Plasma NFL concentration was measured at the Clinical Neurochemistry Laboratory, University of Gothenburg (Mölndal, Sweden) using the NF-Light Advantage kit on a Simoa HD-1 instrument (Quanterix, Billerica, MA, USA)^22,23^.

#### Memory Clinic Cohort, Singapore

Furthermore, we included patients from the Memory Clinic Cohort of the National University Hospital, Singapore (age = 73.98±8.2 years, 51% female), which comprised patients with AD, vascular dementia (VAD), mild cognitive impairment (MCI), vascular cognitive impairment (VCI) and cognitively normal individuals (see Table 1 and Figure 2 for details).

**Table 1.** Demographics of the individuals from the Gothenburg H70 Birth Cohort and Singaporean Memory Clinic Cohort from whom CT datasets were included

| CT datasets |  |  |  |  |  |  |
| --- | --- | --- | --- | --- | --- | --- |
| Variable | H70 Birth Co-<br>hort | Singaporean Memory Clinic Cohort |  |  |  |  |
|  | *n*=917 | AD *n*=73 | MCI *n*=49 | VAD *n*=20 | VCI *n*= 40 | CN *n*=22 |
| Age (years) | 70.44±2.6 | 75.26±1.17 | 76.60±5.97 | 76.55±9.53 | 70.40±10.19 | 68.23±6.5 |
| Gender, female (*n*) | 484 | 48 | 27 | 7 | 10 | 8 |
| Education (years) | 13±4.3 | 4.51±4.38 | 7.208±4.82 | 5.85±3.46 | 6.725±4.17 | 9.52±4.59 |
| ICV (litres) | 1.66±0.21 | 1.8±0.42 | 1.86±0.43 | 1.86±0.39 | 1.52±0.39 | 1.69±0.46 |
| MMSE | 28.9 ± 1.85 | 15.15±4.15 | 22.94±3.69 | 15.10±4.54 | 24.10±3.04 | 27.05±2.08 |
| GM (litres) | 0.77±0.09 | 0.71±0.16 | 0.81±0.19 | 0.69±0.16 | 0.65±0.17 | 0.76±0.21 |
| WM (litres) | 0.59±0.08 | 0.69±0.16 | 0.76±0.15 | 0.65±0.13 | 0.59±0.15 | 0.65±0.16 |
| GM adjusted for CSF | 1.31±0.26 | 0.68±0.09 | 0.77±0.14 | 0.68±0.15 | 0.73±0.12 | 0.83±0.16 |
| GM adjusted for VCSF | 8.1±2.2 | 0.67±0.14 | 0.8±0.18 | 0.69±0.13 | 0.68±0.16 | 0.81±0.21 |
| BV adjusted for CSF | 2.31±0.45 | 1.33±0.17 | 1.48±0.22 | 1.32±0.24 | 1.43±0.21 | 1.56±0.25 |
| BV adjusted for VCSF | 4.58±1.3 | 1.34±0.27 | 1.56±0.32 | 1.33±0.28 | 1.33±0.29 | 1.51±0.36 |

| Biochemical markers |  |  |  |  |  |  |  |
| --- | --- | --- | --- | --- | --- | --- | --- |
| Gothenburg H70 Birth Cohort |  | Singaporean Memory Clinic Cohort |  |  |  |  |  |
| Variable | *n*=316 | Variable | AD *n*=16 | MCI *n*=27 | VAD *n*=10 | VCI *n*= 8 | CN *n*=5 |
| Plasma NfL (pg/mL) | 25.12±25.57 | Plasma NfL (pg/mL) | 36.21±15.14 | 29.85±14.42 | 45.00±35.35 | 28.40±8.84 | 20.32±5.37 |
| CSF NfL (pg/mL) | 906.3±952.5 | Plasma p-Tau 181 (pg/mL) | 18.49±7.82 | 15.94±9.14 | 15.45±5.97 | 17.15±6.73 | 12.24±2.78 |
| CSF p-Tau (pg/L) | 334.04±49.6 | Plasma Aβ 40 (pg/mL) | 305.01±111.9 | 347.02±101.5 | 437.36±206.6 | 295.14±76.02 | 306.85±56.90 |
| CSF Aβ 40 (pg/mL) | 6208.6±1383.47 | Plasma Aβ 42 (pg/mL) | 12.95±5.92 | 12.65±3.85 | 17.74±10.00 | 12.64±2.64 | 12.30±2.76 |
| CSF Aβ 42 (pg/mL) | 715.44±225.559 |  |  |  |  |  |  |

All values are mean ± standard deviation. Abbreviations: Aβ, β-amyloid; AD, Alzheimer's disease; BV, brain volume; CN, cognitively normal; CSF, cerebrospinal fluid; ICV, intracranial volume; MCI, mild cognitive impairment; MMSE, mini-mental state examination; NfL, neurofilament light; p-Tau, phosphorylated tau; VAD, vascular dementia; VCI, vascular cognitive impairment

We included 204 brain CT images (AD, *n*=73; MCI, *n*=49; VAD, *n*=20; VCI, *n*=40; cognitively normal, *n*=22) and 241 MR (AD, *n*=84; MCI, *n*=61; VAD, *n*=25; VCI, *n*=44; cognitively normal, *n*=26) images for our study. For all CT images, paired MR images were available. CT scans were performed using a 256 multislice CT scanner (Philips Medical Systems; slice collimation: 30×0.625mm for CT brain, kVp: 120, mAs: mA modulation with reference mAs of 300) at the National University Hospital, Singapore. MR scans were performed on a Magnetom Trio Tim scanner (3 Tesla; Siemens Healthineers AG, Erlangen, Germany), using a 32-channel head coil, at the Clinical Imaging Research Centre, National University of Singapore.

Demographic data, past medical history, and clinical diagnoses of dementia were obtained. Patients were diagnosed based on results from clinical assessment and neuropsychological tests. Plasma biomarkers (NfL, p-Tau181, Aβ40, and Aβ42) and Aβ status were available for *n*=96/241 participants with available MR images. Blood samples were analysed at the Clinical Neurochemistry Laboratory, University of Gothenburg (Mölndal, Sweden) using a Simoa HD-1 (Quanterix, Billerica, MA, USA) instrument^24^.

### Image processing and analysis

#### Deriving volumetric measures from CT using deep learning models

Previously, we trained U-Net-based deep-learning models to differentiate tissue classes (GM, WM, CSF and ICV) on cranial CT^17^. For the present study, we developed deep-learning models to derive ventricular CSF (VCSF) maps from cranial CT images. Please refer to Supplementary material A1 for the summary of the model development. Altogether five classes were derived from all CT scans using these deep-learning models: GM, WM, CSF, VCSF and ICV. Tissue class volumes were estimated from the deep-learning-derived segmentation maps. Total brain volume (BV) was obtained by summation of GM and WM tissue class volumes. Thus, six atrophy-related volumetric measures and ratios were derived: GM, WM, BV adjusted for CSF, BV adjusted for VCSF, GM adjusted for CSF and GM adjusted for VCSF. The ratios were derived by regressing out the effect of CSF/VCSF from GM/BV^25,26^ (Figure 3).

**Figure 3:**
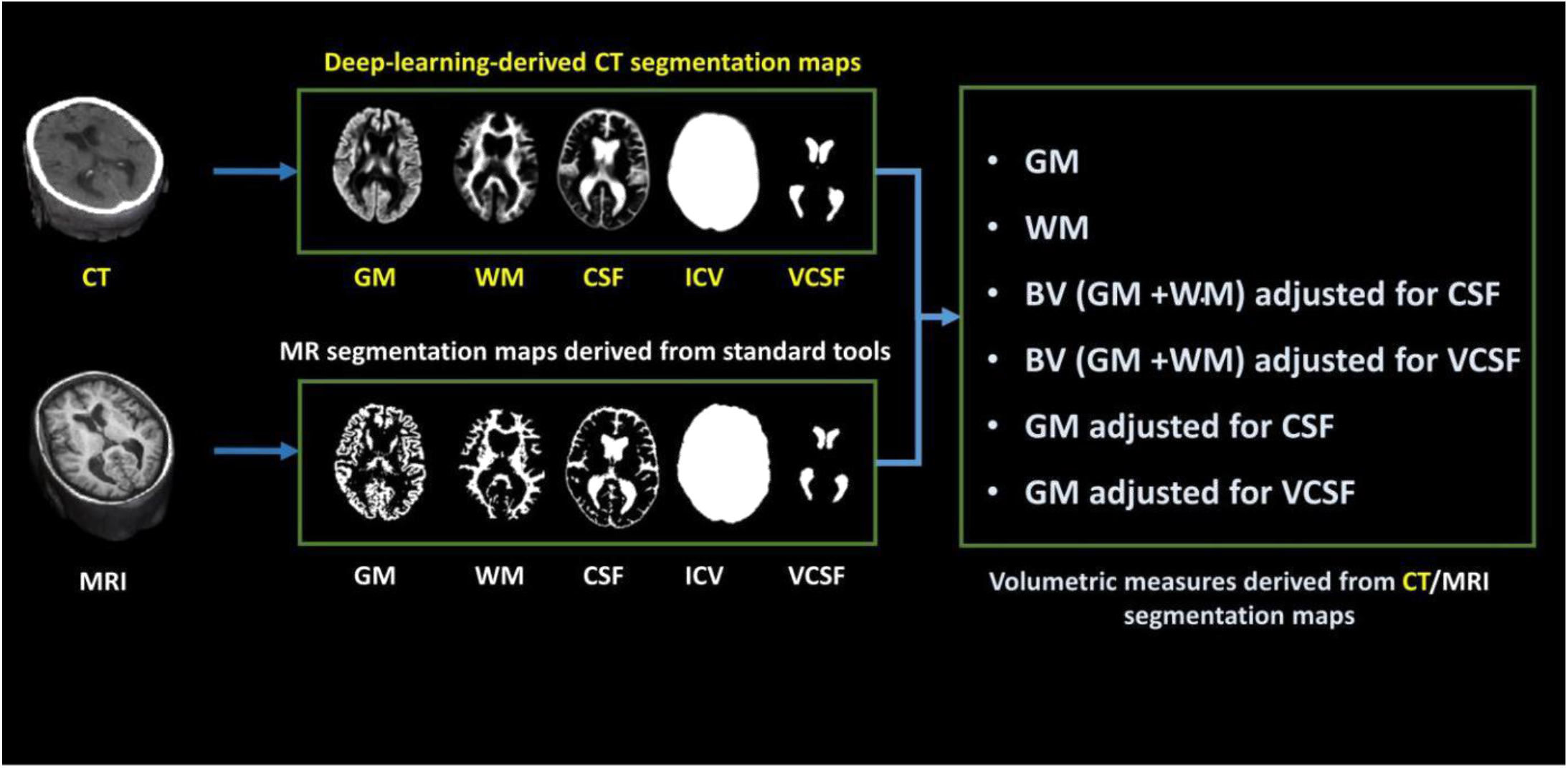
Imaging-derived volumetric measures. The CT-based grey matter (GM), white matter (WM), cerebrospinal fluid (CSF), intracranial volume (ICV) and ventricular cerebrospinal fluid (VCSF) maps were automatically segmented using deep learning techniques. The MR images were automatically segmented to GM, WM, and CSF maps using SPM12, ICV map using Pincram, and VCSF map using MAPER. Six volumetric measures were derived from these segmentation maps. The ratios were obtained by regressing out the effect of the denominator volume from the numerator.

#### Obtaining volumetric measures from MRI

In the Singaporean Memory Clinic Cohort we segmented 241 MRI datasets into GM, WM, and CSF using the unified segmentation routine in SPM12 (https://www.fil.ion.ucl.ac.uk/spm/software/spm12)^27^. VCSF and medial temporal lobe (MTL) volumes were measured using the multi-atlas-based segmentation tool MAPER^28^, and ICV using Pincram^29^. These analyses yielded GM, WM, BV adjusted for CSF, BV adjusted for VCSF, GM adjusted for CSF, GM adjusted for VCSF, and MTL from MR data (Figure 3).

### Statistical analyses

Shapiro–Wilk test was used to examine the Gaussian distribution of the continuous variables in our data (p > 0.05). We compared the volumetric similarity of CT-derived segmentations with MR-derived segmentations in the Singaporean Memory Clinic Cohort using Spearman rank correlation test. The volumetric similarity of CT- and MR-derived segmentations in the Gothenburg H70 Birth Cohort had been tested in our previous work^17^. We further tested associations between the six CT-derived volumetric measures and CSF biomarkers (NfL, p-Tau, Aβ42/40 and Aβ42) and plasma NfL in the Gothenburg H70 Birth Cohort, as well as with plasma biomarkers (NfL, p-Tau181, Aβ42/40 and Aβ42) in the Singaporean Memory Clinic Cohort. In both cohorts, we also tested the relationships of CT-derived volumetric measures with measures of cognition (mini mental state examination (MMSE) and clinical dementia rating sum of boxes (CDR-SB)). Partial spearman correlations between deep-learning-derived CT volumes and other markers of neurodegeneration were employed adjusted for ICV, age, education, and gender. In the Singaporean Memory Clinic Cohort, we compared CT-derived atrophy measures between diagnoses using Kruskal-Wallis tests, and diagnostic accuracy was assessed by measuring the area under the receiver operating characteristic curve (AUC). For comparison in the same cohort, we also tested the relationship between MR-derived volumetric measures and clinical diagnoses, using the same tests and analytic procedures that were used for evaluating CT-based atrophy measures. After conducting the Kruskal-Wallis tests and AUC analysis on MR-based atrophy measures, the diagnostic performance of CT- and MR-based measures was compared. Additionally, to understand the relationship between automatically CT-derived volumes with visual atrophy assessments in the Gothenburg H70 Birth Cohort, we tested the correlation of 1) CT-derived GM with MR-derived GM and MR-MTA, and 2) CT-MTA with MR-MTA and MR-derived GM. The statistical significance threshold was set at p = 0.05. All statistical analyses were performed using IBM SPSS Statistics 27 (IBM Corp, Armonk, NY, U.S.A.) or R version 4.0.3 (2021)^30^.

## 3. Results

Table 1 lists demographic, CT-derived volumetric and biochemical biomarker data for all participants from both cohorts. In the Singaporean Memory Clinic Cohort, mean age and education levels were comparable among diagnostic groups. As expected, AD (15.15±4.15) and VAD (15.10±4.54) patients performed worse on the MMSE than healthy participants (27.05±2.08), and mean plasma NfL levels were higher in AD (36.21±15.14 pg/mL) and VAD (45.00±35.35 pg/mL) compared with cognitively normal individuals (20.32±5.37 pg/mL) and the Gothenburg H70 Birth Cohort participants (25.12±25.57 pg/mL).

In the Singaporean Memory Clinic Cohort, CT volumes correlated strongly with corresponding MR volumes (Spearman’s ρ of 0.93, 0.89 and 0.84 for GM, WM, and CSF tissue classes; Supplementary Figure 1) indicating that our deep-learning-derived CT volumes are comparable to MR volumes derived from established segmentation algorithms. In the Gothenburg H70 Birth Cohort, the inter-modality kappa coefficient between visual ratings of MTA from CT and MR scans performed by a trained rater was 0.40 for the right hemisphere and 0.43 for the left. Importantly, the correlation between CT- and MR-derived GM volumes (ρ=0.872, p<0.001) was stronger than the correlation between CT- and MR-based MTA scores (ρ=0.51, p<0.001) (Supplementary Table 1).

### CT-based atrophy measures across diagnostic groups

In the Singaporean Memory Clinic Cohort, we found that CT-derived deep-learning-based neurodegeneration measurements could distinguish AD patients from cognitively normal subjects (CT-GM adjusted for CSF: AUC=0.88, 95% CI: 0.79, 0.97) and VAD patients from cognitively normal subjects (CT-GM: AUC= 0.91, 95% CI: 0.81, 1.0; Figure 4) yielding AUC values that were comparable to MR-based measurements (AUC for MR-BV adjusted for CSF: AD vs CN: 0.91, 95% CI: 0.85, 0.96; VAD vs CN: 0.91, 95% CI: 0.81, 1.0). CT-derived GM volume measures also yielded comparable AUC values to MR-derived MTL volumes when distinguishing AD patients from normal individuals (CT-GM: AUC= 0.85, MR-MTL: AUC= 0.86; Supplementary Figure 3). Interestingly, CT-derived atrophy measures could distinguish MCI patients (CT-GM adjusted for CSF: AUC= 0.73, 95% CI: 0.62, 0.85) and VCI patients (CT-GM: AUC= 0.7, 95% CI: 0.51, 0.81) from cognitively normal individuals with moderate accuracy (Supplementary Figure 2, Figure 5). The measures mentioned here showed highest AUCs in comparison to other measures. Figure 4, Supplementary Figure 2 and 3 depicts the ROC curves of various measures in distinguishing between different diagnostic groups.

**Figure 4:**
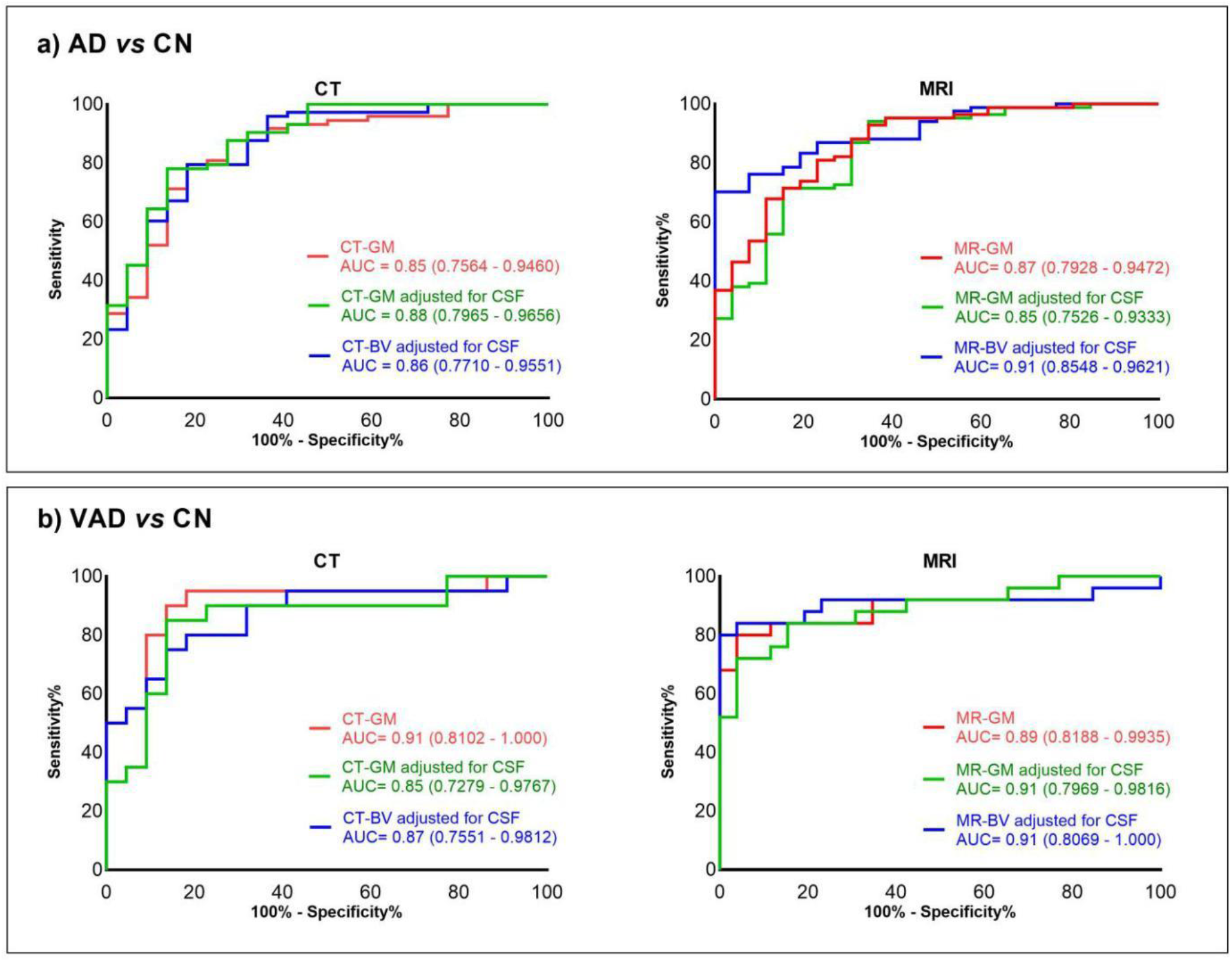
ROC curves for (a) AD vs CN and (b) VAD vs CN using CT- and MR-based atrophy measures in the Memory Clinic Cohort, Singapore (n= 204). All volumes were adjusted for intracranial volume. *Abbreviations: AD, Alzheimer’s disease; AUC, Area under the curve, BV, brain volume; CSF, cerebrospinal fluid; GM, grey matter; CN, cognitively normal; ROC, receiver operating characteristics*

**Figure 5:**
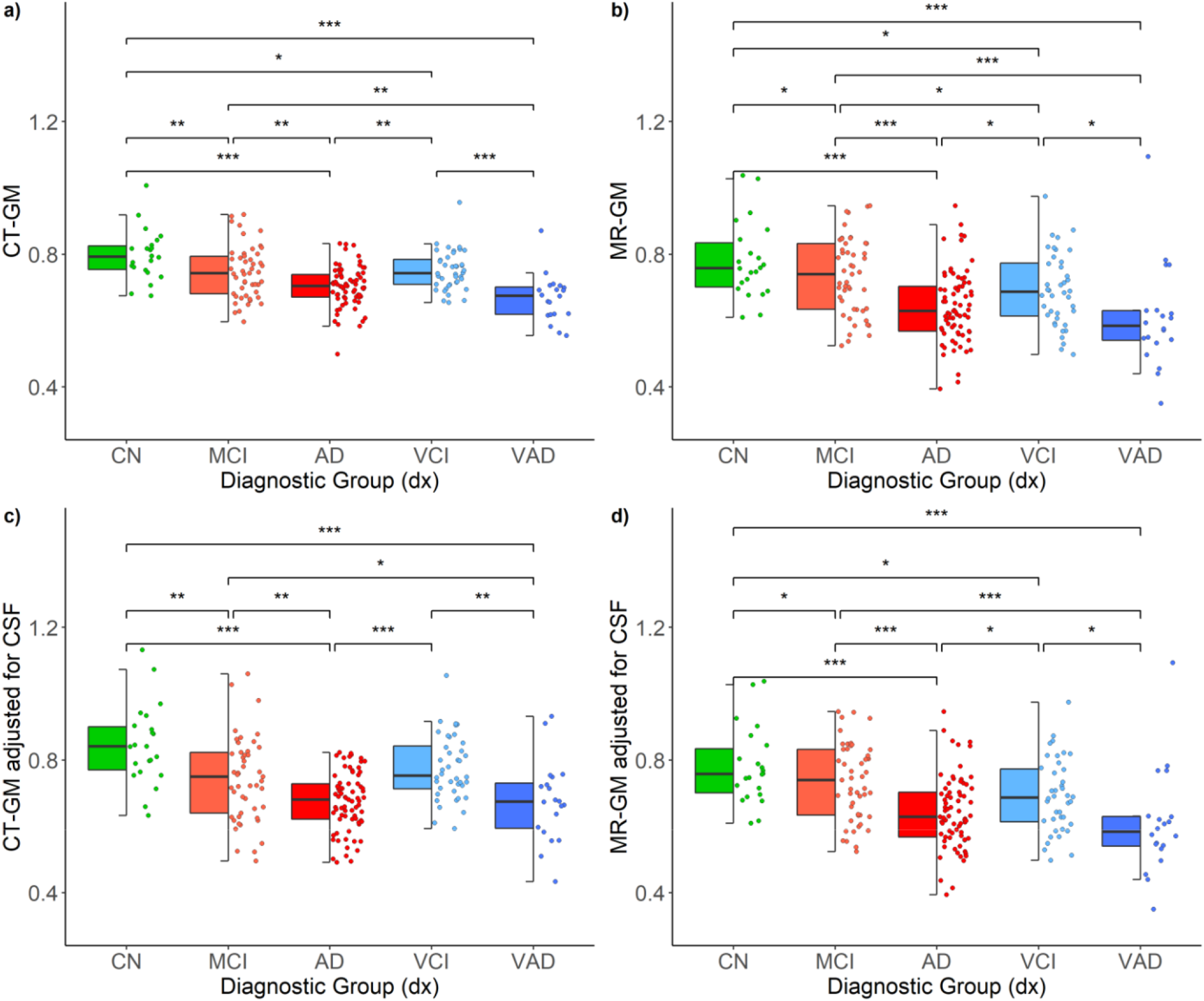
Distribution of CT and MR-derived volumetric measures across diagnostic groups in the Memory Clinic Cohort, Singapore (n=204). ***p<0.001, **p<0.01, *p<0.05; uncorrected p-values derived from Kruskal-Wallis test. All volumes were adjusted for intracranial volume. *Abbreviations: AD, Alzheimer’s disease; BV, brain volume; CN, cognitively normal; CSF, cerebrospinal fluid; GM, grey matter; MCI, mild cognitive impairment; VAD, Vascular dementia; VCI, vascular cognitive impairment*.

Overall, CT-derived volumetric measures differentiated significantly the various diagnostic groups with mean CT-derived volumes being lowest in AD and VAD patients, followed by prodromal dementia groups MCI and VCI and highest in the cognitively normal group (Figure 5). The distribution of CT-derived volumes across various diagnostic groups were comparable to that of MR-derived volumes (Figure 5).

### CT-based atrophy measures and cognition

In the Singaporean Memory Clinic Cohort, deep-learning-derived CT volumes were significantly associated with measures of cognitive impairment as indicated on CDR-SB and MMSE scores (Table 2). Higher CDR-SB and lower MMSE correlated with smaller GM volumes (ρ=-0.44, p<0.001 and ρ= 0.42, p<0.001, respectively) (Figure 6). In the predominantly healthy Gothenburg H70 Birth Cohort, deep-learning-derived CT volumes were significantly, yet more weakly correlated with CDR-SB and MMSE scores (Table 3).

**Table 2.** Summary of correlation analyses in the Singaporean Memory Clinic Cohort

| CT-based measures | Biomarker | $\rho$ | Significance (uncorrected) |
| --- | --- | --- | --- |
| CT-GM | MMSE | 0.42 | p<0.001 |
|  | CDR-SB | -0.44 | p<0.001 |
|  | Plasma NfL | -0.43 | p<0.001 |
|  | Plasma p-tau181 | -0.27 | p=0.03 |
| CT-WM | MMSE | 0.28 | p<0.001 |
|  | CDR-SB | -0.25 | p<0.001 |
|  | Plasma NfL | -0.32 | p=0.008 |
| CT-BV/CSF | MMSE | 0.42 | p<0.001 |
|  | CDR-SB | -0.41 | p<0.001 |
|  | Plasma NfL | -0.4 | p<0.001 |
| | Plasma Aβ42 | 0.25 | p=0.04 |
| CT-GM/CSF | MMSE | 0.37 | p=0.003 |
|  | CDR-SB | -0.37 | p<0.001 |
|  | Plasma NfL | -0.34 | p=0.005 |
| | Plasma Aβ42 | 0.27 | p=0.02 |
| CT-BV/VCSF | MMSE | 0.47 | p<0.001 |
|  | CDR-SB | -0.47 | p<0.001 |
|  | Plasma NfL | -0.49 | p<0.001 |
|  | Plasma p-tau181 | -0.31 | p=0.01 |
| | Plasma Aβ42 | 0.31 | p=0.01 |
| CT-GM/VCSF | MMSE | 0.45 | p<0.001 |
|  | CDR-SB | -0.46 | p<0.001 |
|  | Plasma NfL | -0.41 | p<0.001 |
|  | Plasma p-tau181 | -0.25 | p=0.04 |
| | Plasma Aβ42 | 0.35 | p=0.004 |

Only significant correlations of the partial rank correlation analysis corrected for ICV, age, gender, and education are presented.
Abbreviations: Aβ , $\beta$ -amyloid; BV, brain volume; CDR-SB, clinical dementia rating sum of boxes; CSF, cerebrospinal fluid; GM, grey matter; ICV, intracranial volume; MMSE, mini-mental state examination; NfL, neurofilament light; p-tau, phosphorylated tau; VCSF, ventricular CSF; WM, white matter

**Table 3.**
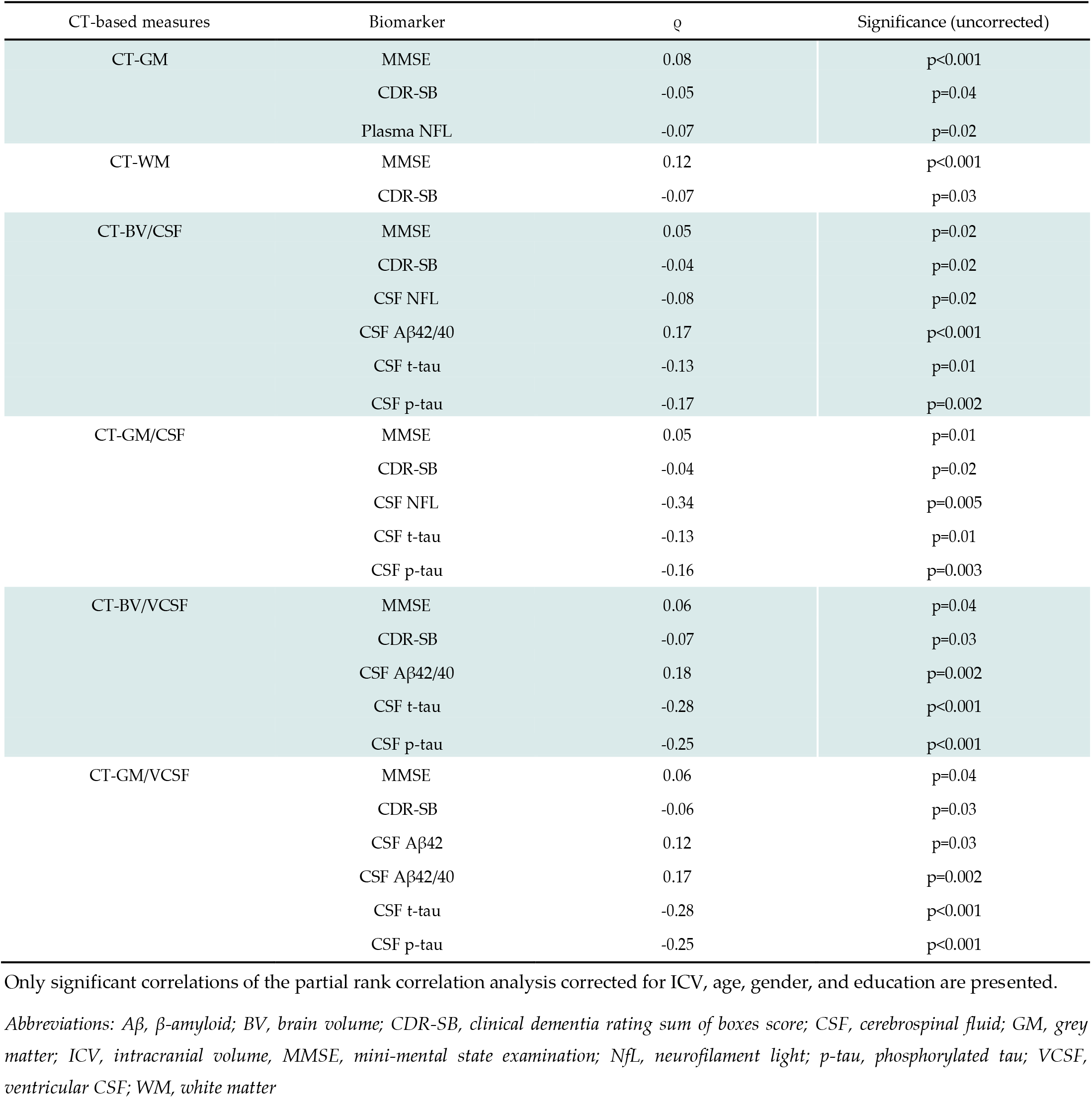
Summary of correlation analyses in the Gothenburg H70 Birth Cohort

**Figure 6:**
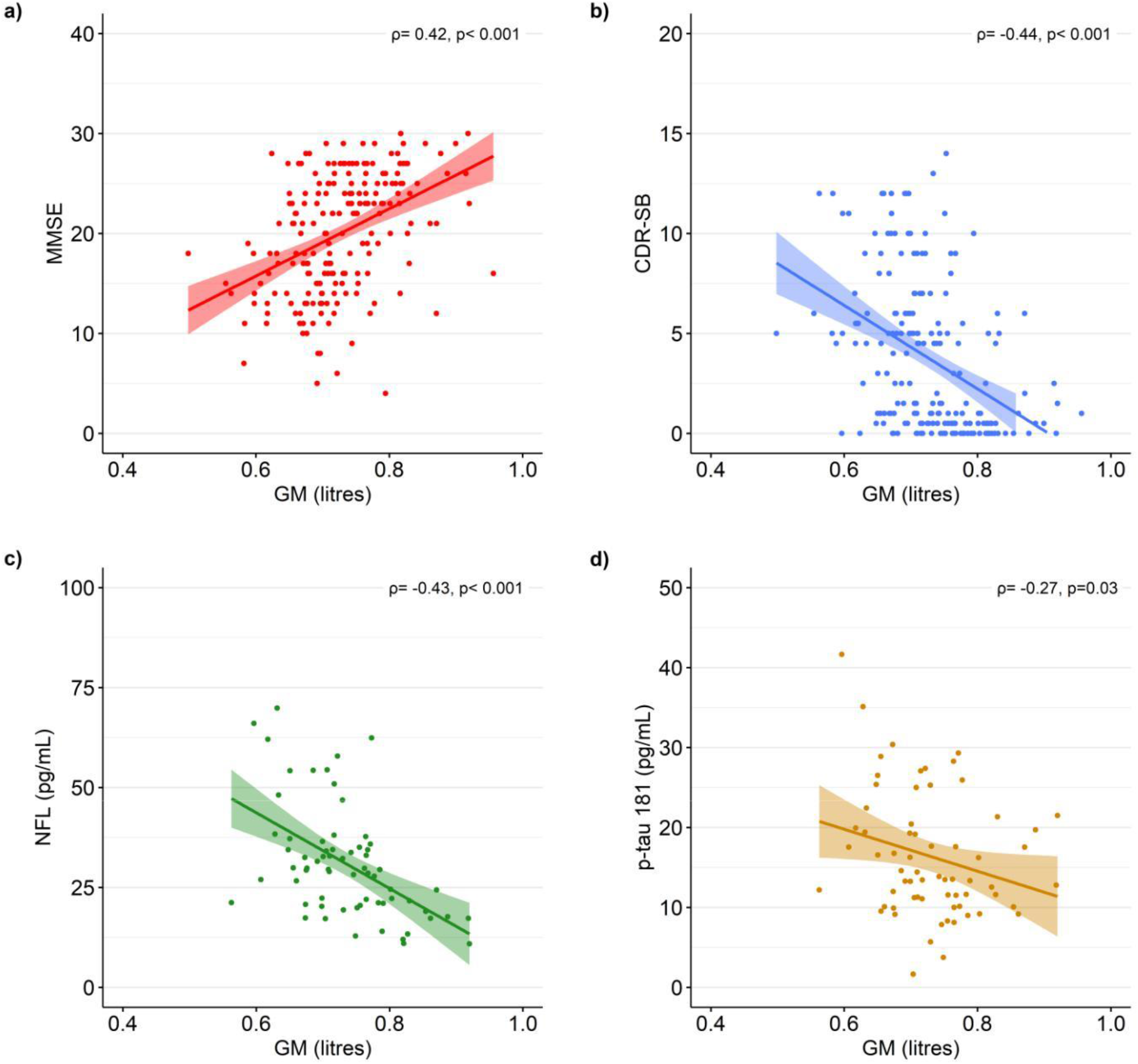
CT-derived volumes with other neurodegenerative disease biomarkers. Correlation of CT-derived GM volumes with a) MMSE, b) CDR-SB, c) plasma NFL, and d) plasma p-tau181 in the Memory Clinic Cohort, Singapore (n=204). The correlation values (ρ) were obtained from partial rank correlation analysis between CT-derived measures and other biomarkers controlled for intracranial volume, age, gender and education. *Abbreviations: CDR-SB, clinical dementia rating sum of boxes; GM, grey matter; MMSE, mini-mental state examination; MR, magnetic resonance; NfL, neurofilament light; p-tau, phosphorylated tau*

### CT-based atrophy measures and their association with biochemical disease markers in plasma and CSF

In the Singaporean Memory Clinic Cohort, CT volumes were significantly associated with plasma-based NfL, p-tau 181, and Aβ42 (Table 2, Figure 6). The strongest correlations were seen with plasma NfL (ρ=-0.49, p<0.001, using CT-BV adjusted for VCSF). In the predominantly healthy Gothenburg H70 Birth Cohort, CT volumes were moderately correlated with plasma levels of NfL (ρ=-0.07, p=0.02, using GM), and CSF levels of NfL, Aβ42, t-tau, p-tau, and notably, Aβ42/40 (ρ=0.18, p=0.002, using CT-BV adjusted for VCSF).

## 4. Discussion

Cranial CT images contain valuable information for characterizing neurodegenerative diseases that can be quantitatively extracted with automatic image analysis. The present study is the first to comprehensively demonstrate that quantitative imaging markers automatically obtained from CT images corroborate the results of standard diagnostic methods used in clinical research and practice. The measurements were enabled by training a dedicated deep neural network with suitable data sets (CT images plus tissue class maps obtained through classical MR image analysis). Once trained, the model works effectively on pure CT data, enabling quantitative assessment of brain atrophy in persons who have not undergone MR scanning. We demonstrated this effectiveness on CT images of 1121 participants in two distinct cohorts, one (*n*=917) and one from individuals consulting a memory clinic (*n*=204). Enabling CT-based assessment of neurodegeneration has important clinical potential as CT scanners are more widely available than MR equipment, and patients with contraindications for MR scanning could still be examined with CT.

We investigated the association of deep-learning-derived CT-based atrophy measures with established markers of neurodegenerative diseases. Our main findings are that brain atrophy measures from our model (1) differentiate between patients and healthy controls, with similar performance as MR-based measures, and (2) strongly correlate with relevant cognitive, biochemical and neuroimaging markers of neurodegenerative diseases associated with dementia. Together, these findings indicate that deep-learning-derived CT-based atrophy measures are promising imaging markers for the first-line assessment of neurodegenerative diseases.

The first key finding of this study is that deep-learning-derived CT-based atrophy measures differentiated between various diagnostic groups with high accuracy, notably between cognitively normal and patients with cognitive severity. The distribution pattern of CT-derived atrophy measures across various diagnostic groups is consistent with findings from previous studies^13,31,32^. In the MCI and VCI groups, we measured volumes that were larger than those of the AD and VAD groups, but smaller than those of the cognitively normal group. An interesting finding is that CT-derived atrophy measures differentiated between MCI and cognitively normal groups (AUC: 0.73, Supplementary Figure 2, Figure 5), indicating a potential application in the early detection of dementia and neurodegeneration. Among the six atrophy measures derived from the five direct CT volumetric measures, GM, GM adjusted for CSF, and BV adjusted for CSF showed the best performance in differentiating between diagnostic groups.

The second key finding of our study is that deep-learning-derived CT-based atrophy measures are associated with cognition and biochemical markers of neurodegenerative disorders. In both cohorts, lower CT-derived volumes were associated with lower MMSE, and higher levels of CDR-SB also correlated with lower levels of CT-derived volumes. Other studies have shown that higher levels of plasma and CSF NfL correlate with lower MMSE scores, decreased brain volume, and reduced cortical thickness^33,34^. We found significant correlations between higher levels of plasma NfL with lower levels of CT-derived volumes. In the Gothenburg H70 Birth Cohort, higher levels of CSF NfL were correlated with lower levels of CT-derived volumetric measures. Studies by Karikari et al.^35^, Moscoso et al.^36^, and Wang et al.^37^ showed that higher plasma p-Tau181 correlates with smaller brain volumes. In the Singaporean Memory Clinic Cohort, higher levels of plasma p-Tau181 were correlated with lower CT-derived volumes. Together, our findings suggest that CT-derived atrophy measures correlate well with clinical and biochemical measures. Among the six deep-learning-derived CT-based atrophy measures, GM, GM adjusted for CSF, BV adjusted for VCSF, and GM adjusted for VCSF correlated with most biochemical markers of neurodegeneration in the Singaporean Memory Clinic Cohort (Table 2), as did GM adjusted for VCSF, GM adjusted for CSF, BV adjusted for VCSF and BV adjusted for CSF in the Gothenburg H70 Birth Cohort (Table 3).

We further found that CT-derived atrophy measures correlate strongly with MR-based measures and display comparable diagnostic performance, notably also compared with MR-MTL (Supplementary Figure 3), the most established quantitative measure of medial temporal lobe atrophy^38–40^. Atrophy of the MTL is an important diagnostic biomarker of many neurodegenerative disorders, most specifically in AD^38,41,42^. Further, we found that the intermodality agreement between our automated CT- and established MRI-segmentations was greatly improved compared to the agreement between visual CT and MRI assessment. In the Gothenburg H70 Birth Cohort, the intermodality kappa coefficient between visual ratings of MTA from CT and MR scans performed by a trained rater was 0.40 for the right hemisphere and 0.43 for the left hemisphere, and the correlation between CT-MTA and MR-MTA was ρ=0.51. The volumetric correlation between CT-GM and MR-GM derived from our automated and established MR-based methods was ρ=0.87 for the same datasets. Currently, brain atrophy in CT is predominantly evaluated using visual assessments, while for differential diagnosis MRI is preferred. Our findings emphasise the potential of automated CT analysis for the evaluation of brain atrophy at the first-line examination of cognitive decline, with faster and more reproducible results than visual ratings, while showing comparable performance to MRI. Our deep learning models perform tissue classification on head CT images in under one minute.

Strengths of our study include that we investigated the association of CT-derived markers with established and novel markers of neurodegenerative diseases (blood- and CSF-based as well as cognitive) in two large well-characterised cohorts for which paired CT and MRI data were available. The first cohort predominantly consisted of cognitively normal participants, and the other consisted of various diagnosis groups of dementia. This allowed us to train our deep learning models on the predominantly healthy cohort and validate the models on both cohorts. By having access to both CT and MR scans of the same subjects, we were able to validate the structural similarity of CT-derived segmentation maps and compare the diagnostic performance of CT-based atrophy measures in both cohorts.

Interestingly, the deep learning models developed in our previous study17 worked well on the Singaporean Memory Clinic Cohort without reference to MRI data and without further training. One of the widely discussed limitations of deep learning models is the lack of reproducibility^43^. Our study shows the transferability of the trained models to previously unseen datasets acquired on other scanners. Limitations of our study include that we had CSF markers, amyloid status, and plasma markers available for 96 out of 241 participants in the Memory Clinic Cohort. Hence the clinical diagnoses of patients reported in this study are likely a reflection of cognitive status and associated underlying pathology rather than diagnoses based on biomarker evidence of a specific aetiology. Additionally, our deep learning models were trained on MR-derived labels only, therefore our study cannot answer whether alternative means of generating training labels on CT images (manual or other) would have been beneficial. Further, we explored the clinical applications of deep-learning-derived CT-based atrophy measures on cross-sectional data. The diagnostic performance of CT-derived atrophy measures on longitudinal data is yet to be explored. In the future, we plan to conduct prospective studies to study the impact of longitudinal effects of neurodegeneration on CT-derived atrophy measures.

In summary, deep-learning-derived CT-based volumetric measures are associated with relevant imaging, cognitive, and biochemical markers of neurodegenerative diseases. These volumetric measures differentiate between patients with dementia diagnoses, even at early disease stages, and cognitively healthy individuals, offering comparable diagnostic performance to established MR-based markers of neurodegenerative diseases. We propose our automated method to produce CT-derived volumetric measures that can support clinical dementia diagnostics, even in early disease stages. Together with novel blood biomarkers, CT-derived atrophy measures will provide highly useful tools for the first-line examination of individuals presenting with cognitive decline.

## Funding

J.B.P. is supported by grants from the Center for Medical Innovation, the Swedish Research Council, Neurotech, Alzheimerfonden, Hjärnfonden, a Senior Researcher Position grant, Gamla Tjänarinnor, Demensfonden and Stohnes grants. HZ is a Wallenberg Scholar supported by grants from the Swedish Research Council (#2018-02532), the European Union’s Horizon Europe research and innovation programme under grant agreement No 101053962, Swedish State Support for Clinical Research (#ALFGBG-71320), the Alzheimer Drug Discovery Foundation (ADDF), USA (#201809-2016862), the AD Strategic Fund and the Alzheimer’s Association (#ADSF-21-831376-C, #ADSF-21-831381-C, and #ADSF-21-831377-C), the Bluefield Project, the Olav Thon Foundation, the Erling-Persson Family Foundation, Stiftelsen för Gamla Tjänarinnor, Hjärnfonden, Sweden (#FO2022-0270), the European Union’s Horizon 2020 research and innovation programme under the Marie Skłodowska-Curie grant agreement No 860197 (MIRIADE), the European Union Joint Programme – Neurodegenerative Disease Research (JPND2021-00694), and the UK Dementia Research Institute at UCL (UKDRI-1003). KB is supported by the Swedish Research Council (#2017-00915), the Alzheimer Drug Discovery Foundation (ADDF), USA (#RDAPB-201809-2016615), the Swedish Alzheimer Foundation (#AF-742881), Hjärnfonden, Sweden (#FO2017-0243), the Swedish state under the agreement between the Swedish government and the County Councils, the ALF-agreement (#ALFGBG-715986), and European Union Joint Program for Neurodegenerative Disorders (JPND2019-466-236). KB is supported by the Swedish Research Council (#2017-00915), the Alzheimer Drug Discovery Foundation (ADDF), USA (#RDAPB-201809-2016615), the Swedish Alzheimer Foundation (#AF-930351, #AF-939721 and #AF-968270), Hjärnfonden, Sweden (#FO2017-0243 and #ALZ2022-0006), the Swedish state under the agreement between the Swedish government and the County Councils, the ALF-agreement (#ALFGBG-715986 and #ALFGBG-965240), the European Union Joint Program for Neurodegenerative Disorders (JPND2019-466-236), the National Institute of Health (NIH), USA, (grant #1R01AG068398-01), and the Alzheimer’s Association 2021 Zenith Award (ZEN-21-848495). SK was financed by grants from the Swedish state under the agreement between the Swedish government and the county councils, the ALF-agreement (ALFGBG-965923, ALFGBG-81392, ALF GBG-771071). The Alzheimerfonden (AF-842471, AF-737641, AF-939825). The Swedish Research Council (2019-02075), Psykiatriska Forskningsfonden, Stiftelsen Demensfonden, Stiftelsen Hjalmar Svenssons Forskningsfond, Stiftelsen Wilhelm och Martina Lundgrens vetenskapsfond. MS is supported by the Knut and Alice Wallenberg Foundation (Wallenberg Centre for Molecular and Translational Medicine; KAW 2014.0363), the Swedish Research Council (#2017-02869), the Swedish state under the agreement between the Swedish government and the County Councils, the ALF-agreement (#ALFGBG-813971), and the Swedish Alzheimer Foundation (#AF-740191). Image analysis computations were in part carried out with resources provided by the Swedish National Infrastructure for Computing (SNIC), partially funded by the Swedish Research Council through grant agreement no. 2018-05973. Gothenburg H70 Birth Cohort was financed by grants from the Swedish state under the agreement between the Swedish government and the county councils, the ALF-agreement (ALF965812, ALF 716681), the Swedish Research Council (2012-5041, 2015-02830, 2013-8717, 2017-00639, 2019-01096, 2022-00882), Swedish Research Council for Health, Working Life and Wellfare (2013-1202, 2018-00471, AGECAP 2013-2300, 2013-2496, 2018-00471),Konung Gustaf V:s och Drottning Victorias Frimurarestiftelse, Hjärnfonden (FO2014-0207, FO2016-0214, FO2018-0214, FO2019-0163, FO2020-0235), Alzheimerfonden (AF-554461, AF-647651, AF-743701, AF-844671, AF-930868, AF-940139, AF-968441), Eivind och Elsa K:son Sylvans stiftelse

## Data Availability

The Gothenburg H70 Birth cohort and Memory Clinic Cohort, Singapore cannot openly share data according to existing ethical and data sharing approvals, however, relevant data can and will be shared with research groups after submitting a research proposal which has to be approved by the respective study coordinators.

## Ethics Approval

The Memory Clinic Cohort Study, Singapore (study protocol number DEM4333) was approved by the National Healthcare Group Domain Specific Review Board (Reference number: NHG DSRB 2018/01098-SRF0004). The H70 study was approved by the Regional Ethical Review Board in Gothenburg (Approval Numbers: 869-13, T076-14, T166-14, 976-13, 127-14, T936-15, 006-14, T703-14, 006-14, T201-17, T915-14, 959-15, T139-15), and by the Radiation Protection Committee (Approval Number: 13-64).

## Supplementary material

### A1 Development of deep learning models for VCSF segmentation from head CT

For model development, we used 744 paired CT and MR images from the predominantly cognitively normal Gothenburg H70 Birth Cohort. We pre-processed all paired CT and MR images using SPM12 (http://www.fil.ion.ucl.ac.uk/spm), running on MATLAB 2020a. We segmented MR images into VCSF labels using MAPER. To represent the CT images and MR labels in a common image matrix, MR images were coregistered to their paired CT images using SPM12. The coregistration between each CT-MR pair was visually assessed and 10 pairs were excluded due to faulty coregistration. The pre-processed datasets consisting of CT images and their paired, co-registered MR labels were subdivided into training and cross-validation groups. The datasets were randomly split to perform three-fold cross-validation training. For each model, one-fold was held out as the unseen test dataset, and two folds were further grouped into 400 training and 100 validation datasets. We developed and trained the 2D U-Net models using the procedures described in Srikrishna et al., 2021. Briefly, we trained 2D U-Net based deep learning models to differentiate between various tissue classes in the head CT. The segmentation patterns were studied from their paired MR labels. U-Net based deep learning models were created and trained to accept CT scans and MR labels as input. The U-Net was designed to perform slice wise processing of input data where each input image was processed as a stack of 2D slices with size 512 x 512 pixels. In total, 12,000 training slices and 3,000 validation slices were used to train 1,177,649 trainable hyper-parameters. The inputs were fed into the model, and learning was executed using the Keras module. The batch size was 16. Callback features were used such as early stopping and automatic reduction of learning rate with respect to rate of training. The model was trained for 50 epochs with 750 samples per epoch in approximately 540 mins. We trained the U-Nets also on an Nvidia GeForce RTX 2080 Ti GPU using TensorFlow 2.0 and Keras 2.3.1.

**Supplementary Figure 1.**
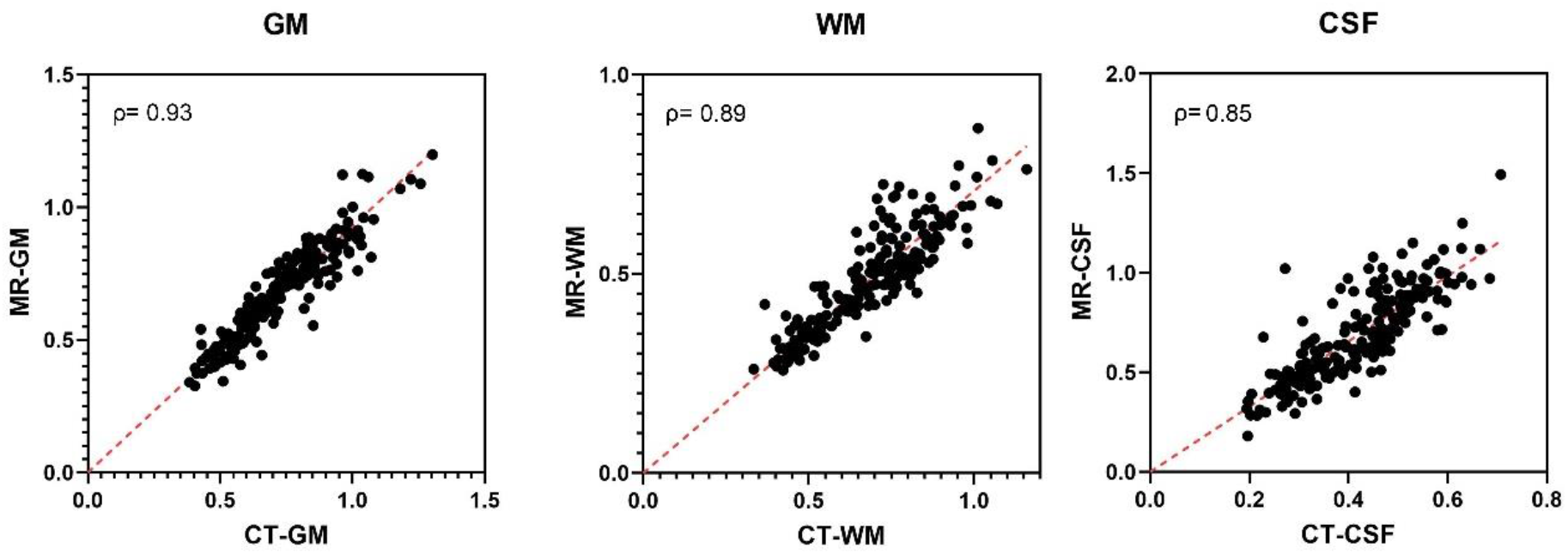
Correlation between deep-learning-derived segmentations from unseen test CT scans and MR scans of Singaporean Memory Clinic Cohort (*n*= 204). *Abbreviations: CSF, cerebrospinal fluid; GM, grey matter; WM, white matter*

**Supplementary Figure 2.**
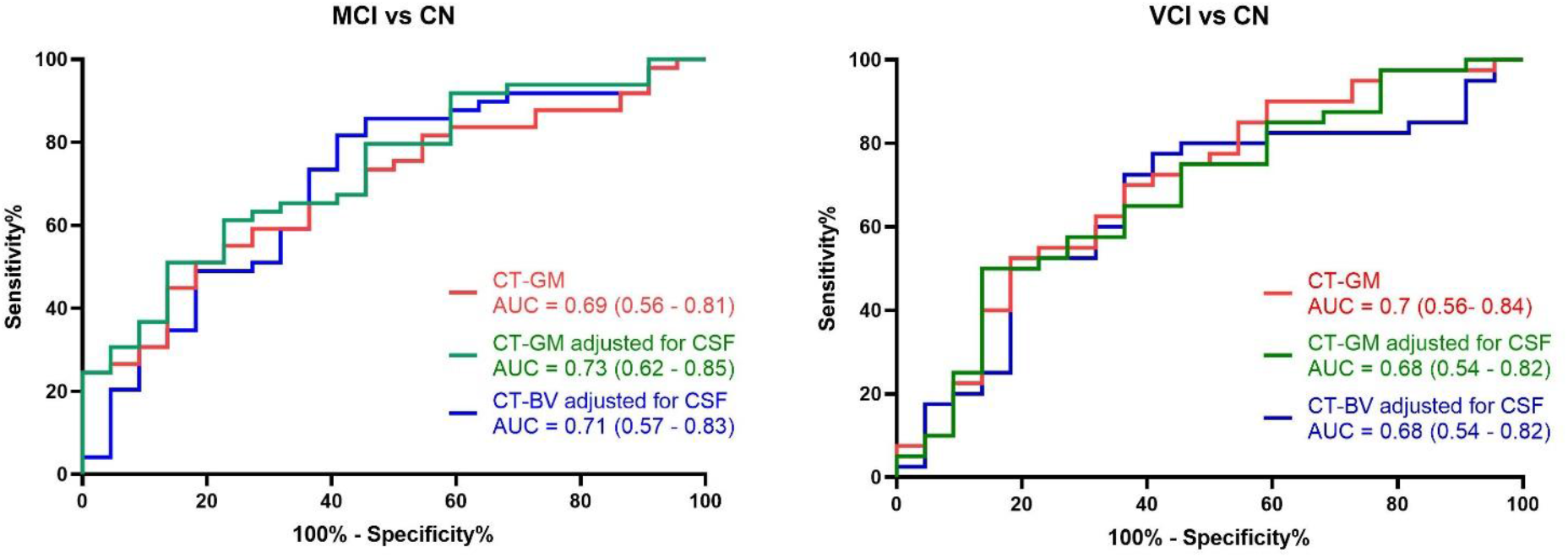
ROC curves for MCI vs CN (a) and VCI vs CN (b) for CT-based atrophy measures in the Singaporean Memory Clinic Cohort. *Abbreviations: AUC, Area under the curve; BV, brain volume; CSF, cerebrospinal fluid; GM, grey matter; CN, cognitively normal; MCI, mild cognitive impairment; ROC, receiver operating characteristic; VCI, vascular contributions of cognitive impairment*.

**Supplementary Table 1.**
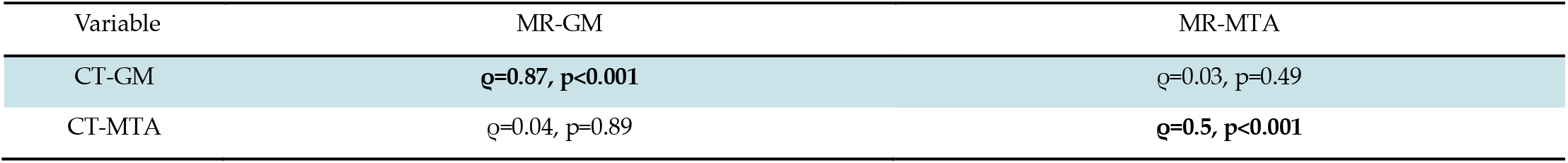
Association of CT-derived GM and MTA with MR-derived GM and MTA. Spearman’s correlation coefficient (ρ) measures the strength and direction of association between volumetric measures and MTA scores from Gothenburg H70 Birth Cohort.

| Variable | MR-GM | MR-MTA |
| --- | --- | --- |
| CT-GM | $\rho=0.87, p<0.001$ | $\rho=0.03, p=0.49$ |
| CT-MTA | $\rho=0.04, p=0.89$ | $\rho=0.5, p<0.001$ |

**Supplementary Figure 3.**
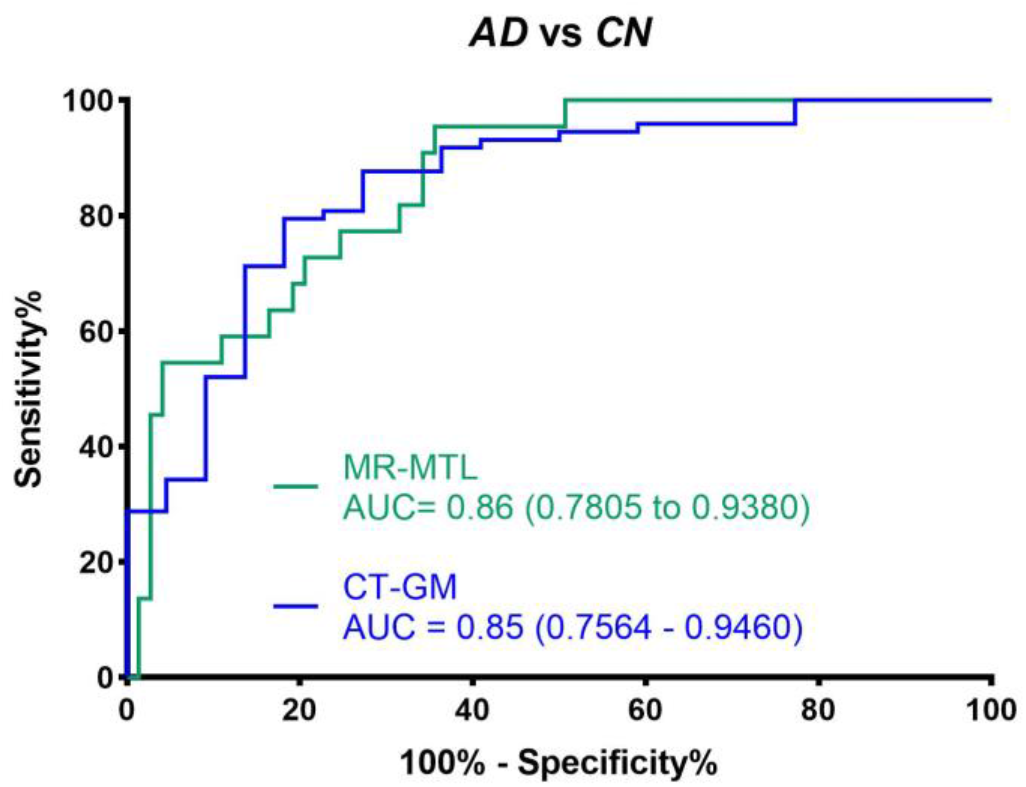
ROC curves for AD vs CN for CT-GM and MR-MTL of Singaporean Memory Clinic Cohort. *Abbreviations: AD, Alzheimer’s disease; AUC, Area under the curve; GM, grey matter; CN, cognitively normal; MTL, medial temporal lobe; ROC, receiver operating characteristic*.

## Notes

### Competing Interest Statement

KB has served as a consultant, at advisory boards, or at data monitoring committees for Abcam, Axon, BioArctic, Biogen, JOMDD/Shimadzu. Julius Clinical, Lilly, MagQu, Novartis, Ono Pharma, Pharmatrophix, Prothena, Roche Diagnostics, and Siemens Healthineers, and is a co-founder of Brain Biomarker Solutions in Gothenburg AB (BBS), which is a part of the GU Ventures Incubator Program, outside the work presented in this paper. SK has served at scientific advisory boards and / or as consultant for Geras Solutions and Biogen. MS has served at an advisory board for Servier Pharmaceuticals, outside of the present work. HZ has served at scientific advisory boards and/or as a consultant for Abbvie, Acumen, Alector, Alzinova, ALZPath, Annexon, Apellis, Artery Therapeutics, AZTherapies, CogRx, Denali, Eisai, Nervgen, Novo Nordisk, Passage Bio, Pinteon Therapeutics, Prothena, Red Abbey Labs, reMYND, Roche, Samumed, Siemens Healthineers, Triplet Therapeutics, and Wave, has given lectures in symposia sponsored by Cellectricon, Fujirebio, Alzecure, Biogen, and Roche, and is a co-founder of Brain Biomarker Solutions in Gothenburg AB (BBS), which is a part of the GU Ventures Incubator Program (outside submitted work).

### Funding Statement

HZ is a Wallenberg Scholar supported by grants from the Swedish Research Council (#2018-02532), the European Research Council (#681712), Swedish State Support for Clinical Research (#ALFGBG-720931), the Alzheimer Drug Discovery Foundation (ADDF), USA (#201809-2016862), the AD Strategic Fund and the Alzheimers Association (#ADSF-21-831376-C, #ADSF-21-831381-C and #ADSF-21-831377-C), the Olav Thon Foundation, the Erling-Persson Family Foundation, Stiftelsen f ör Gamla Tjänarinnor, Hjärnfonden, Sweden (#FO2019-0228), the European Unions Horizon 2020 research and innovation programme under the Marie Skłodowska-Curie grant agreement No 860197 (MIRIADE), and the UK Dementia Research Institute at UCL. KB is supported by the Swedish Research Council (#2017-00915), the Alzheimer Drug Discovery Foundation (ADDF), USA (#RDAPB-201809-2016615), the Swedish Alzheimer Foundation (#AF-742881), Hjärnfonden, Sweden (#FO2017-0243), the Swedish state under the agreement between the Swedish government and the County Councils, the ALF-agreement (#ALFGBG-715986), and European Union Joint Program for Neurodegenerative Disorders (JPND2019-466-236). KB is supported by the Swedish Research Council (#2017-00915), the Alzheimer Drug Discovery Foundation (ADDF), USA (#RDAPB-201809-2016615), the Swedish Alzheimer Foundation (#AF-930351, #AF-939721 and #AF-968270), Hjärnfonden, Sweden (#FO2017-0243 and #ALZ2022-0006), the Swedish state under the agreement between the Swedish government and the County Councils, the ALF-agreement (#ALFGBG-715986 and #ALFGBG-965240), the European Union Joint Program for Neurodegenerative Disorders (JPND2019-466-236), the National Institute of Health (NIH), USA, (grant #1R01AG068398-01), and the Alzheimers Association 2021 Zenith Award (ZEN-21-848495). SK was financed by grants from the Swedish state under the agreement between the Swedish government and the county councils, the ALF-agreement (ALFGBG-965923, ALFGBG-81392, ALF GBG-771071). The Alzheimerfonden (AF-842471, AF-737641, AF-15939825). The Swedish Research Council (2019-02075), Psykiatriska Forskningsfonden, Stiftelsen Demensfonden, Stiftelsen Hjalmar Svenssons Forskningsfond, Stiftelsen Wilhelm och Martina Lundgrens vetenskapsfond. MS is supported by the Knut and Alice Wallenberg Foundation (Wallenberg Centre for Molecular and Translational Medicine; KAW 2014.0363), the Swedish Research Council (#2017-02869), the Swedish state under the agreement between the Swedish government and the County Councils, the ALF-agreement (#ALFGBG-813971), and the Swedish Alzheimer Foundation (#AF-740191). Image analysis computations were in part carried out with resources provided by the Swedish National Infrastructure for Computing (SNIC), partially funded by the Swedish Research Council through grant agreement no. 2018-05973.

## References

1. Ross CA, Poirier MA. Protein aggregation and neurodegenerative disease. Nature Medicine. 2004;10(7):S10. doi:10.1038/nm1066

2. Jack CR, Bennett DA, Blennow K, et al. NIA-AA Research Framework: Toward a biological definition of Alzheimer’s disease. Alzheimer’s & Dementia. 2018;14(4):535–562. doi:10.1016/j.jalz.2018.02.018

3. Parnetti L, Eusebi P. Cerebrospinal fluid biomarkers in Alzheimer’s disease: an invaluable tool for clinical diagnosis and trial enrichment. Journal of Alzheimer’s Disease. 2018;64(1):S281–S287. doi:10.3233/JAD-179910

4. Pini L, Pievani M, Bocchetta M, et al. Brain atrophy in Alzheimer’s disease and aging. Ageing research reviews. 2016;30:25–48. doi:10.1016/j.arr.2016.01.002

5. Frisoni GB, Fox NC, Jack CR, Scheltens P, Thompson PM. The clinical use of structural MRI in Alzheimer disease. Nat Rev Neurol. 2010;6(2):67–77. doi:10.1038/nrneurol.2009.215

6. Ehrenberg AJ, Khatun A, Coomans E, et al. Relevance of biomarkers across different neurodegenerative diseases. Alz Res Therapy. 2020;12(1):56. doi:10.1186/s13195-020-00601-w

7. Young PNE, Estarellas M, Coomans E, et al. Imaging Biomarkers in Neurodegeneration: Current and Future Practices. Vol 12. BioMed Central Ltd.; 2020. doi:10.1186/s13195-020-00612-7

8. Ashton NJ, Hye A, Rajkumar AP, et al. An update on blood-based biomarkers for non-Alzheimer neurodegenerative disorders. Nat Rev Neurol. 2020;16(5):265–284. doi:10.1038/s41582-020-0348-0

9. Leuzy A, Cullen NC, Mattsson-Carlgren N, Hansson O. Current advances in plasma and cerebrospinal fluid biomarkers in Alzheimer’s disease. Current Opinion in Neurology. 2021;34(2):266–274. doi:10.1097/WCO.0000000000000904

10. Wattjes MP, Henneman WJ, van der Flier WM, et al. Diagnostic imaging of patients in a memory clinic: comparison of MR imaging and 64–detector row CT. Radiology. 2009;253(1):174–183. doi:10.1148/radiol.2531082262

11. Pasi M, Poggesi A, Pantoni L. The use of CT in dementia. International psychogeriatrics. 2011;23(S2):S6-S12. doi:10.1017/S1041610211000950

12. Sluimer JD, van der Flier WM, Karas GB, et al. Whole-brain atrophy rate and cognitive decline: longitudinal MR study of memory clinic patients. Radiology. 2008;248(2):590–598. doi:10.1148/radiol.2482070938

13. Orellana C, Ferreira D, Muehlboeck JS, et al. Measuring Global Brain Atrophy with the Brain Volume/Cerebrospinal Fluid Index: Normative Values, Cut-Offs and Clinical Associations. Neurodegener Dis. 2016;16(1-2):77–86. doi:10.1159/0004424433

14. Wei M, Shi J, Ni J, et al. A new age-related cutoff of medial temporal atrophy scale on MRI improving the diagnostic accuracy of neurodegeneration due to Alzheimer’s disease in a Chinese population. BMC Geriatrics. 2019;19(1):59. doi:10.1186/s12877-019-1072-8

15. Aguilar C, Edholm K, Simmons A, et al. Automated CT-based segmentation and quantification of total intracranial volume. European radiology. 2015;25(11):3151–3160. doi:Multiclass brain tissue segmentation in 4d ct using convolutional neural networks

16. Van De Leemput SC, Meijs M, Patel A, Meijer FJ, Van Ginneken B, Manniesing R. Multiclass brain tissue segmentation in 4d ct using convolutional neural networks. IEEE Access. 2019;7:51557–51569. doi:srikrishna

17. Srikrishna M, Pereira JB, Heckemann RA, et al. Deep learning from MRI-derived labels enables automatic brain tissue classification on human brain CT. NeuroImage. 2021;244:118606. doi:10.1016/j.neuroimage.2021.118606

18. Ronneberger O, Fischer P, Brox T. U-net: Convolutional networks for biomedical image segmentation. In: Springer; 2015:234–241. doi:10.48550/arXiv.1505.04597

19. Rydberg Sterner T, Ahlner F, Blennow K, et al. The Gothenburg H70 Birth cohort study 2014–16: design, methods and study population. European journal of epidemiology. 2019;34(2):191–209. doi:10.1007/s10654-018-0459-8

20. Lindberg O, Kern S, Skoog J, et al. Effects of amyloid pathology and the APOE ε4 allele on the association between cerebrospinal fluid Aβ38 and Aβ40 and brain morphology in cognitively normal 70-years-olds. Neurobiol Aging. 2021;101:1–12. doi:10.1016/j.neurobiolaging.2020.10.033

21. Skoog I, Kern S, Najar J, et al. A Non-APOE Polygenic Risk Score for Alzheimer’s Disease Is Associated With Cerebrospinal Fluid Neurofilament Light in a Representative Sample of Cognitively Unimpaired 70-Year Olds. J Gerontol A Biol Sci Med Sci. 2021;76(6):983–990. doi:10.1093/gerona/glab030

22. Dittrich A, Ashton NJ, Zetterberg H, et al. Plasma and CSF NfL are differentially associated with biomarker evidence of neurodegeneration in a community-based sample of 70-year-olds. Alzheimer’s & Dementia: Diagnosis, Assessment & Disease Monitoring. 2022;14(1):e12295. doi:10.1002/dad2.12295

23. Benedet AL, Leuzy A, Pascoal TA, et al. Stage-specific links between plasma neurofilament light and imaging biomarkers of Alzheimer’s disease. Brain. 2020;143(12):3793–3804. doi:10.1093/brain/awaa342

24. Janelidze S, Mattsson N, Palmqvist S, et al. Plasma P-tau181 in Alzheimer’s disease: relationship to other biomarkers, differential diagnosis, neuropathology and longitudinal progression to Alzheimer’s dementia. Nat Med. 2020;26(3):379–386. doi:10.1038/s41591-020-0755-1

25. Voevodskaya O, Simmons A, NordenskjÃ\Pld R, et al. The effects of intracranial volume adjustment approaches on multiple regional MRI volumes in healthy aging and Alzheimer’s disease. Frontiers in Aging Neuroscience. 2014;6(OCT):264. doi:10.3389/fnagi.2014.00264

26. O’Brien LM, Ziegler DA, Deutsch CK, Frazier JA, Herbert MR, Locascio JJ. Statistical adjustments for brain size in volumetric neuroimaging studies: Some practical implications in methods. Psychiatry Res. 2011;193(2):113–122. doi:10.1016/j.pscychresns.2011.01.007

27. Ashburner J, Friston KJ. Unified segmentation. Neuroimage. 2005;26(3):839–851. doi:10.1016/j.neuroimage.2005.02.018

28. Heckemann RA, Keihaninejad S, Aljabar P, et al. Improving intersubject image registration using tissue-class information benefits robustness and accuracy of multi-atlas based anatomical segmentation. Neuroimage. 2010;51(1):221–227. doi:10.1016/j.neuroimage.2010.01.072

29. Heckemann RA, Ledig C, Gray KR, et al. Brain extraction using label propagation and group agreement: Pincram. PloS one. 2015;10(7):e0129211. doi:10.1371/journal.pone.0129211

30. Core R. Team. R: A Language and Environment for Statistical Computing, 2015. Published online 2021.

31. Chen W, Song X, Zhang Y, et al. An MRI-based semiquantitative index for the evaluation of brain atrophy and lesions in Alzheimer’s disease, mild cognitive impairment and normal aging. Dement Geriatr Cogn Disord. 2010;30(2):121–130. doi:10.1159/000319537

32. Spulber G, Simmons A, Muehlboeck JS, et al. An MRI-based index to measure the severity of Alzheimer’s disease-like structural pattern in subjects with mild cognitive impairment. J Intern Med. 2013;273(4):396–409. doi:10.1111/joim.12028

33. Pereira JB, Westman E, Hansson O. Association between cerebrospinal fluid and plasma neurodegeneration biomarkers with brain atrophy in Alzheimer’s disease. Neurobiology of Aging. 2017;58:14–29. doi:10.1016/j.neurobiolaging.2017.06.002

34. Zetterberg H, Skillbäck T, Mattsson N, et al. Association of cerebrospinal fluid neurofilament light concentration with Alzheimer disease progression. JAMA Neurology. 2016;73(1):60–67. doi:10.1001/jamaneurol.2015.3037

35. Karikari TK, Pascoal TA, Ashton NJ, et al. Blood phosphorylated tau 181 as a biomarker for Alzheimer’s disease: a diagnostic performance and prediction modelling study using data from four prospective cohorts. The Lancet Neurology. 2020;19(5):422–433. doi:10.1016/S1474-4422(20)30071-5

36. Moscoso A, Grothe MJ, Ashton NJ, et al. Longitudinal associations of blood phosphorylated Tau181 and neurofilament light chain with neurodegeneration in Alzheimer disease. JAMA neurology. 2021;78(4):396–406.

37. Wang YL, Chen J, Du ZL, et al. Plasma p-tau181 Level Predicts Neurodegeneration and Progression to Alzheimer’s Dementia: A Longitudinal Study. Front Neurol. 2021;12:695696. doi:10.3389/fneur.2021.695696

38. Korf ESC, Wahlund LO, Visser PJ, Scheltens P. Medial temporal lobe atrophy on MRI predicts dementia in patients with mild cognitive impairment. Neurology. 2004;63(1):94–100. doi:10.1212/01.WNL.0000133114.92694.93

39. Jack CR, Petersen RC, Xu YC, et al. Medial temporal atrophy on MRI in normal aging and very mild Alzheimer’s disease. Neurology. 1997;49(3):786–794. doi:10.1212/wnl.49.3.786

40. Cotta Ramusino M, Altomare D, Bacchin R, et al. Medial temporal lobe atrophy and posterior atrophy scales normative values. NeuroImage: Clinical. 2019;24:101936. doi:10.1016/j.nicl.2019.101936

41. Westman E, Cavallin L, Muehlboeck JS, et al. Sensitivity and specificity of medial temporal lobe visual ratings and multivariate regional MRI classification in Alzheimer’s disease. PloS one. 2011;6(7):e22506.

42. Wahlund LO, Westman E, van Westen D, et al. Imaging biomarkers of dementia: recommended visual rating scales with teaching cases. Insights into imaging. 2017;8(1):79–90.

43. Beam AL, Manrai AK, Ghassemi M. Challenges to the reproducibility of machine learning models in health care. Jama. 2020;323(4):305–306.

